# Azithromycin mass drug administration to reduce child mortality in Niger (AVENIR II): a master protocol for a cluster-randomized adaptive platform trial to evaluate community-based health interventions

**DOI:** 10.1101/2025.06.17.25329431

**Authors:** Ahmed M. Arzika, Abdou Amza, Sani Ousmane, Ramatou Maliki, Ibrahim Almou, Nasser Galo, Nasser Harouna, Alio Mankara, Bawa Aichatou, Ousseini Boubacar, Elodie Lebas, Brittany Peterson, Carolyn Brandt, Andrea Picariello, Angela Cheng, Travis C. Porco, Thuy Doan, Benjamin F. Arnold, Thomas M. Lietman, Kieran S. O’Brien

## Abstract

**Background:** Trials have demonstrated that azithromycin mass drug administration (MDA) to children 1-59 months old reduces mortality, but increases antimicrobial resistance (AMR). The World Health Organization recommends that programs include mortality and AMR monitoring. Niger is expanding the azithromycin MDA for child survival program nationwide.

**Methods:** To establish program monitoring and leverage the infrastructure to evaluate other community health interventions, AVENIR II is designed as a cluster-randomized adaptive platform trial with monitoring and re-randomization every 2 years. The initial focus is to monitor under-5 mortality, AMR, implementation, and safety as the program expands in Niger. All eligible primary health center catchment areas (Centre de Santé Intégrés, CSIs) will be included in biannual oral azithromycin MDA to children 1-59 months old. A subset will be randomized to delay MDA for the first 2 years, after which they will receive MDA and another subset will be randomized to stop MDA for the next 2 years. The proportion randomized to delay or stop will be determined using an adaptive algorithm including: 1) results of prior azithromycin MDA mortality trials, 2) expert opinion on the appropriate ethical balance between delivering the program and monitoring AMR, and 3) statistical power to detect a programmatically relevant difference between arms. We anticipate 5-10% of CSIs will be randomized to delay or stop at each randomization. Mortality and AMR will be monitored at baseline and every 2 years. Implementation and safety outcomes will be monitored continuously. To enable ongoing monitoring while ensuring program access, CSIs receiving MDA will be re-randomized using the adaptive algorithm updated with new mortality results and no CSI will go without MDA for more than 2 years. In this platform design, additional arms may be added or dropped based on information from other studies, updates to guidelines, or preferences of Niger policymakers, and other interventions may be evaluated.

**Discussion:** The risk of AMR has led to caution in the implementation of azithromycin MDA. We present a design that enables continued rigorous evaluation of program impact on key outcomes, with flexibility to evaluate other interventions as well.

**Trial registration:** clinicaltrials.gov (NCT06358872), registered April 2024

## BACKGROUND

Several cluster-randomized trials have demonstrated that biannual azithromycin mass drug administration (MDA) to children 1-59 months old reduces child mortality by 14-18% in West Africa.(1–3) As under-5 mortality has remained persistently high in this setting, this approach may complement existing child survival interventions and health systems strengthening efforts. Although azithromycin MDA is also associated with increases in antimicrobial resistance (AMR), the World Health Organization (WHO) determined that the immediate child survival benefits outweigh the uncertain future AMR risks.(4) The WHO’s conditional guidelines for azithromycin MDA focus on implementation in high mortality sub-Saharan African settings.(4) The guidelines recommend targeting 1-11-month-olds instead of 1-59-month-olds to reduce antibiotic use, with continued monitoring of mortality, AMR, and adverse events to support updates to the risk-benefit assessment.(4) The recent AVENIR I trial confirmed that treating all children 1-59 months of age reduced mortality and was unable to demonstrate that targeting only children 1-11 months of age was effective.(3) In fact, the trial provided evidence that treating all 1-59 - month-olds provided indirect benefits to 1-11-month-olds in addition to direct benefits – 1-11-month mortality was 17% lower in communities receiving 1-59-month MDA compared to those receiving 1-11-month MDA alone.(3)

Along with several other West African countries, Niger is expanding azithromycin MDA for child survival as a national program.(5) Given evidence from the AVENIR I trial, the program will include all children 1-59 months of age. A Ministry of Health (MOH) committee has been established to oversee the design and implementation of the program as well as to determine the approach to monitoring mortality, AMR, implementation outcomes, and safety over time. The committee determined that the risk of AMR necessitated a rigorous monitoring design that would allow for evaluation of the causal impact of this program on mortality and resistance. This enables the committee to make evidence-based decisions about continuing and expanding the program or stopping.

Here we describe the protocol for an adaptive platform trial that was designed to rigorously monitor this program while flexibly adjusting to changing priorities over time.(6) The design involves the progressive national expansion of the program in Niger with active monitoring of mortality, AMR, implementation, and safety outcomes in a subset of communities using a randomized design. This approach allows us to 1) monitor the real-world effectiveness of this program in reducing child mortality over time as the epidemiologic landscape shifts, 2) attribute the causal impact of the program on AMR, and 3) add arms over time to evaluate other interventions, leveraging the investment in the program and monitoring infrastructure as community health needs change.

## METHODS/DESIGN

### Study Design Overview and Specific Aims

AVENIR II is a cluster-randomized adaptive platform trial to evaluate community health interventions in Niger with monitoring and re-randomization approximately every 2 years. The initial objective is to monitor under-5 mortality and AMR as the azithromycin MDA for child survival program expands throughout Niger. All eligible primary health center catchment areas (Centre de Santé Intégrés, CSIs) will be included in biannual oral azithromycin MDA to children 1-59 months old. A subset of CSIs will be randomized to delay MDA for the first 2 years, after which they will receive MDA and another subset will be randomized to stop MDA for the next 2 years (Figure 1). This results in two trials: Trial 1 (effectiveness) compares azithromycin MDA to delayed treatment over the first 2 years, and Trial 2 (stopping) compares continuing azithromycin MDA to stopping treatment over the second 2 years. Trial 1 enables the MOH committee to determine the initial impact of the program on mortality and AMR to decide whether the program should continue expanding. Trial 2 allows for an evaluation of mortality and AMR when stopping the program after 2 years versus continuing. Currently, no guidelines exist on how long to continue the program and this trial will provide initial evidence to support decision-making around stopping. The proportion randomized to delay or stop will be determined using an adaptive algorithm. We anticipate that 5-10% of CSIs will be randomized to delay or stop at each randomization. Mortality and AMR will be monitored at baseline and every 2 years. Implementation outcomes and adverse events will be monitored continuously. To enable continued monitoring while ensuring access to the program, CSIs receiving MDA will be re-randomized using the adaptive algorithm updated with new mortality results and no CSI will go without MDA for more than 2 years.

**Figure 1.**
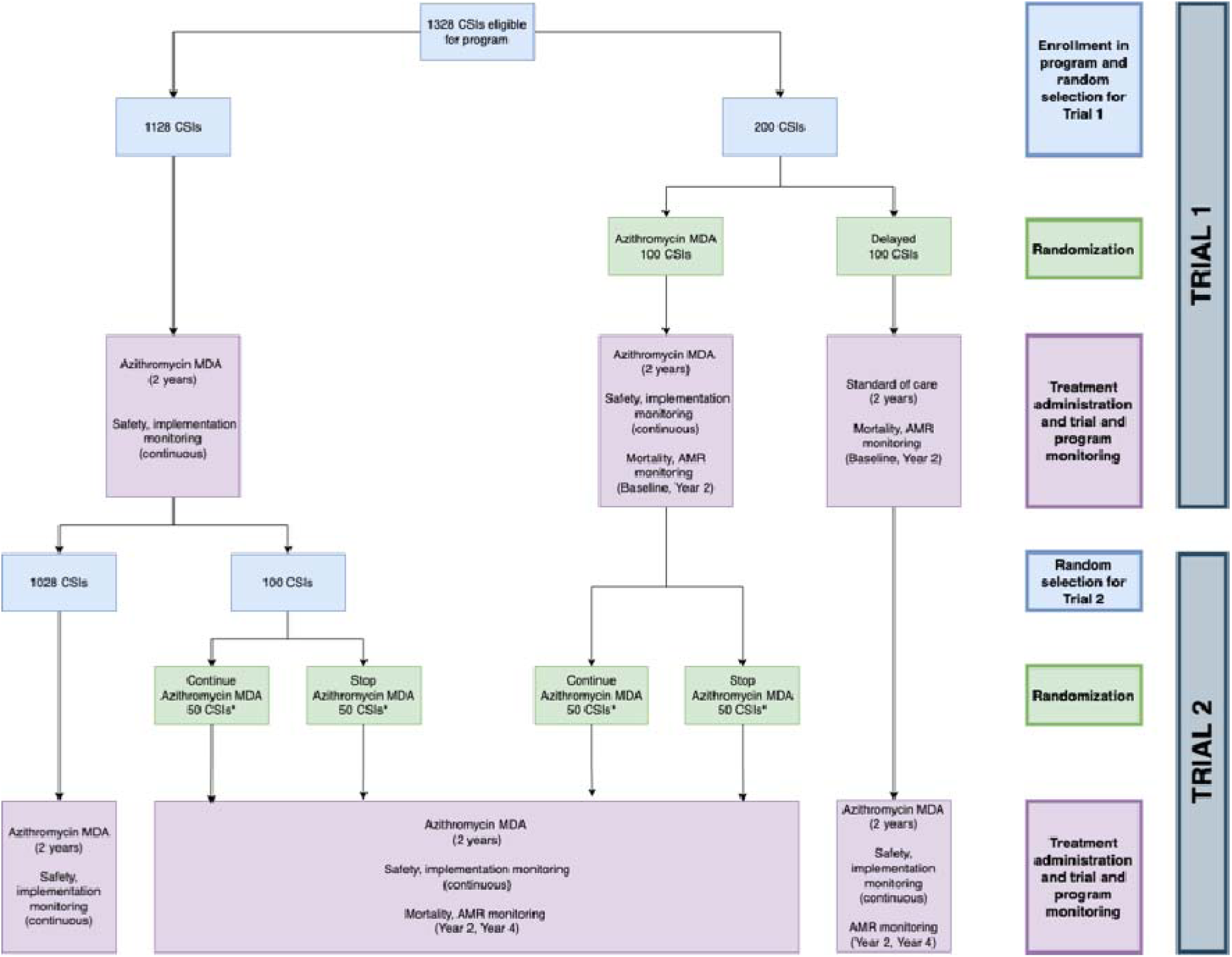
Trial Design Overview. Flow diagram of national program, Trial 1 (effectiveness) and Trial 2 (stopping). Blue boxes describe enrollment and random selection, green boxes describe randomization, and pink boxes describe intervention and monitoring activities.

In this platform design, a key feature is flexibility without loss of rigor. We aim to assess program performance under conditions that may necessitate addition or removal of study arms, or changes in timing of implementation between regions. Additional treatment arms may be added to the trial or arms may be dropped from the trial based on additional information from other studies, updates to the WHO guidelines, or preferences of programs or policymakers in Niger. Potential interventions to be added to the trial will be decided by the MOH committee and study team. With such changes, protocol details, power calculations, and analysis plans will be updated accordingly in the protocol, sub-protocols, and statistical analysis plan. New treatment arms are ideally added at the time of re-randomization every 2 years, though they can be added at other times. New treatment arms will be described in sub-protocols referencing this master protocol.

The initial design includes the following specific aims:

**Specific Aim 1 - Mortality:** to evaluate the effectiveness of azithromycin MDA to reduce under- 5 mortality every 2 years in a real-world program setting.

a. Trial 1: In the first 2 years of the trial, we will compare under-5 mortality in CSIs randomized to azithromycin MDA or delayed treatment.
b. Trial 2: In the second 2 years of the trial, we will compare under-5 mortality in CSIs randomized to stop or continue azithromycin MDA.

### Specific Aim 2 – Antimicrobial resistance

a. To compare the prevalence of macrolide-resistant pneumococcus from nasopharyngeal swabs in population-based samples of children 1–59 months old every 2 years.
i. Trial 1: In the first 2 years of the trial, we will compare the prevalence of macrolide resistance in CSIs randomized to azithromycin MDA or delayed treatment.
ii. Trial 2: In the second 2 years of the trial, we will compare the prevalence of macrolide resistance in CSIs randomized to stop or continue azithromycin MDA.
iii. At Year 4, we will compare the prevalence of macrolide resistance in CSIs randomized to have had 4 years of azithromycin MDA or 2 years (continue vs original delayed then treated)
b. To compare the load of genetic determinants of resistance to macrolides from rectal swabs in children 1–59 months old every 2 years.
i. Trial 1: In the first 2 years of the trial, we will compare the load of macrolide resistance determinants in CSIs randomized to azithromycin MDA or delayed treatment.
ii. Trial 2: In the second 2 years of the trial, we will compare the load of macrolide resistance determinants in CSIs randomized to stop or continue azithromycin MDA.
iii. At Year 4, we will compare the load of macrolide resistance determinants in communities randomized to have had 4 years of azithromycin MDA or 2 years (continue vs original delayed then treated)

### Specific Aim 3 – Implementation

a. To evaluate equity of treatment coverage and identify barriers, facilitators, and implementation strategies to increase coverage.
b. To map the intervention delivery process and identify the flow of inputs required to achieve high treatment coverage.
c. To determine program costs, cost-effectiveness, and variations in implementation strategies that affect costs.
d. To monitor fidelity of the administration of the intervention to the protocol.

This protocol was prepared according to SPIRIT reporting guidelines.(7)

### Setting, Timeline, and Participants

All accessible and secure regions in Niger will be progressively included in the program over a 2-year period. Program activities will begin in the Dosso, Tahoua, and Maradi regions in mid-2024, in the Zinder region at the end of 2024, and in the remaining regions in 2025 and 2026 (Figure 2). The first 4 regions launching in 2024 are expected to encompass approximately 75% of the target population and will be included in the trials and monitoring activities. We anticipate reaching approximately 3.3 million children and delivering 15.4 million doses of azithromycin over the 4-year period in the overall program (Table 1).

**Figure 2.**
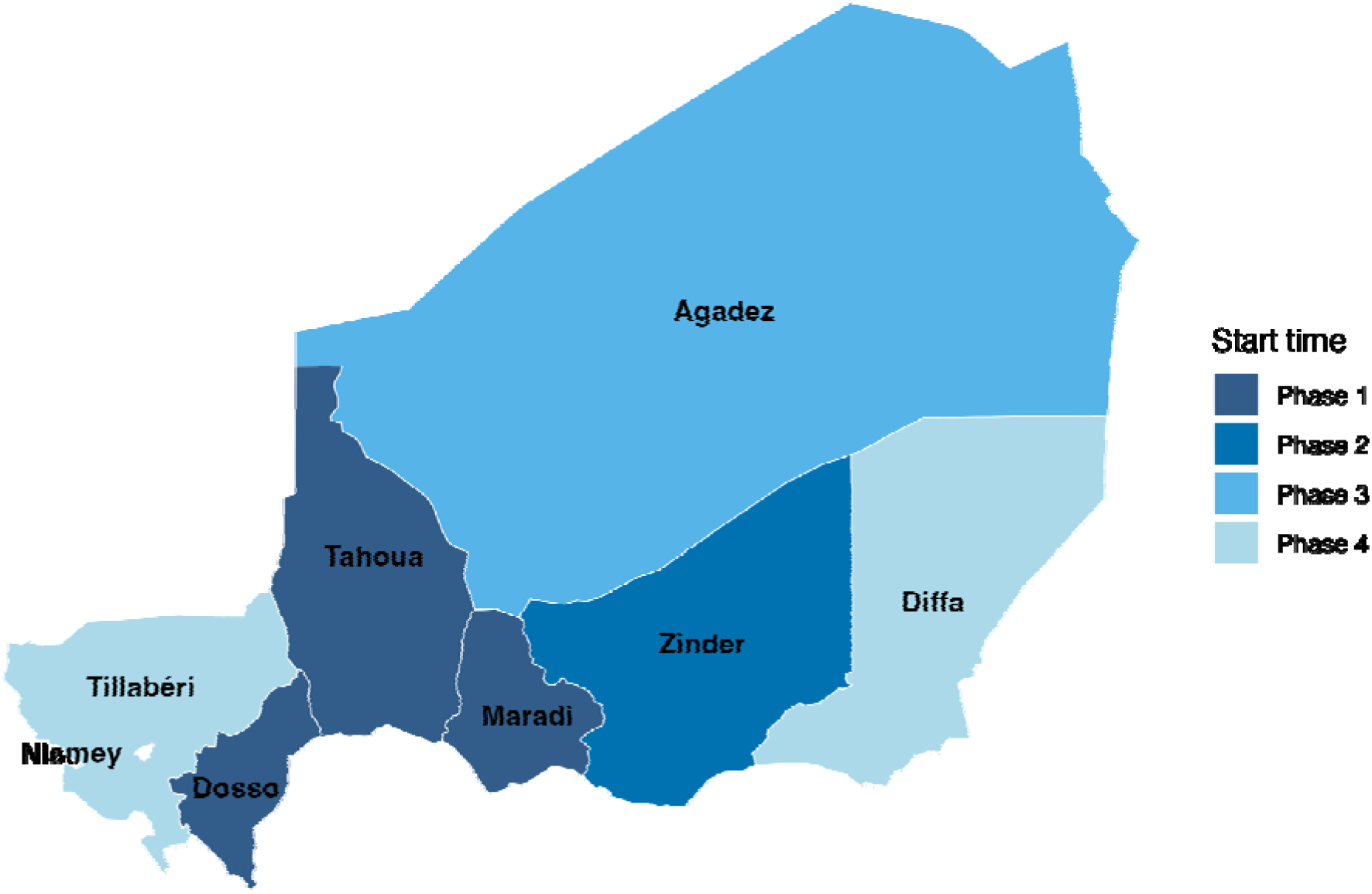
Implementation Rollout Plan. Map of the regions of Niger colored according to the progressive scale-up plan. Borders indicate the 7 regions in Niger plus the capital of Niamey, which is not included in the scale-up. All regions are included in the national program and the Dosso, Tahoua, Maradi, and Zinder regions are included in the trial. Phase 1 is expected to begin in the second and third quarters of 2024. Phase 2 is expected to begin in the fourth quarter of 2024. Phase 3 is expected to begin in the second quarter of 2025. and Phase 4 is expected to begin in the second quarter of 2026.

**Table 1.** Timeline and impact of progressive national scale-up of azithromycin mass administration in Niger. Phase 1 is expected to begin in the second and third quarters of 2024. Phase 2 is expected to begin in the fourth quarter of 2024. Phase 3 is expected to begin in the second quarter of 2025. and Phase 4 is expected to begin in the second quarter of 2026.

| Region | Trial or Program | Start Time | Districts | CSIs | MDAs<sup>1</sup> | Estimated eligible children 1-59m<sup>2</sup> | Estimated number of delivered overall<sup>3</sup> |
| --- | --- | --- | --- | --- | --- | --- | --- |
| Dosso | Both | Phase 1 | 8 | 192 | 7 | 431,461 | 2,416,182 |
| Tahoua | Both | Phase 1 | 13 | 267 | 7 | 670,002 | 3,752,011 |
| Maradi | Both | Phase 1 | 9 | 216 | 7 | 665,826 | 3,728,626 |
| Zinder | Both | Phase 2 | 11 | 219 | 6 | 731,986 | 3,513,533 |
| Agadez | Program only | Phase 3 | 7 | 92 | 5 | 67,298 | 269,192 |
| Tillabéri | Program only | Phase 4 | 13 | 270 | 3 | 585,215 | 1,404,516 |
| Diffa | Program only | Phase 4 | 6 | 72 | 3 | 129,642 | 311,141 |
| **Total** |  |  | **67** | **1328** | **38** | **3,281,430** | **15,395,200** |

CSI, Centre de Santé Intégré (primary health center); m, months; MDA, mass drug administration
<sup>1</sup> Assuming twice yearly MDA
<sup>2</sup> Population estimates including rural and peri-urban communities, produced by the Institute for Disease Modeling (IDM)
<sup>3</sup> Using IDM population estimates and assuming 80% coverage

Eligibility for the program is considered at the CSI and individual levels. CSI-level eligibility for MDA includes location in a participating region that is safe and accessible for study and/or program teams, is designated as primarily rural by the study team, and has verbal consent from community leaders. Child-level eligibility for MDA includes residence in the catchment area of a participating CSI, age between 1 and 59 months old, and verbal informed consent from the child’s caregiver. CSIs will be randomly selected for inclusion in trial and outcome monitoring activities as described below.

### Randomization, Adaptation, and Masking

*Randomization unit and monitoring sample.* The randomization unit for this trial is the CSI, which is a primary health care center with an average catchment area of approximately 20 communities (*grappes)*, each of which include an average of approximately 100 children under 5. A random sample of 200 CSIs eligible for the program will be selected for participation in the trials as a stratified sample of 50 CSIs from each of the four regions launching the program in year 1: Dosso, Tahoua, Maradi, and Zinder. Trial 1 will include this initial sample of 200 CSIs, which will be randomized 1:1 to receive the intervention or to delayed treatment. After 2 years, the 100 delayed CSIs will be treated by the program and not included in Trial 2. The 100 treated CSIs will be included in Trial 2, along with an additional 100 randomly selected CSIs from the larger program, again stratified by region. Note that the samples sizes included in Trial 2 are subject to the adaptation as described in the next section.

*Adaptation.* Allocation probabilities for AVENIR II will adapt from the AVENIR I trial based on 1-59 month mortality using the following algorithm.(3) Given all outcome measurements available through the primary endpoint for AVENIR, we will fit a negative binomial model to the data from the 1-59 month and placebo arms. From the model, we will estimate each arm’s log mortality rate and its standard error. We will then draw 10,000 replicates of log mortality rates from the estimated distributions, assuming they are normally distributed, and in each draw of the joint distribution we will determine which arm has the lowest mortality rate. We will estimate the probability that each arm has the lowest mortality rate as the proportion of 10,000 replicates in which the arm has the lowest mortality rate. In AVENIR II, there will be no tempering of allocation probabilities because we have sufficient information to be protected from uninformed swings in allocation.

The adaptive algorithm will include a minimum probability for the delayed and stopping arms to enable continued comparison to untreated groups while ensuring the majority retain access to treatment based on statistical power and expert opinion. The preliminary value is 0.10, which was used in the AVENIR I algorithm, and confirmed by a survey of expert opinion. As we anticipate 894 total CSIs in Dosso, Tahoua, Maradi, and Zinder regions, a 10% allocation would represent 90 CSIs in the delayed group, or approximately 23 CSIs per region (rounded to whole numbers). To account for potential exclusions due to accessibility, we will include 25 delayed CSIs per region with an equal number of treated CSIs per region, for a total of 50 CSIs per region and 200 CSIs in the monitoring population of the trials. (Figure 1). The adaptation will be updated after Trial 1 to inform Trial 2. Allocation sequences will be generated by the UCSF biostatistician in R (R Foundation for Statistical Computing).

*Masking.* Given the nature of the intervention, participants, community health workers (CHWs) delivering the intervention, and team members supervising the program will not be masked. One biostatistician and one data analyst will remain unmasked to prepare the randomization sequence. Masked personnel include outcome assessors as well as the biostatistician and data analyst conducting the data analyses.

### Interventions

Sensitization campaigns will be conducted at the community, district, regional, and national levels before program and study activities begin. Leaders at these levels will continue to be engaged by the study team throughout the course of the trial and in the dissemination of results. All children 1–59 months of age will be recruited for the intervention. CHWs will visit every household in eligible communities in a door-to-door fashion every 6 months and will obtain verbal informed consent from caregivers of eligible children before administering treatment.

Azithromycin will be administered biannually as a single, directly observed dose of 20 mg/kg (up to the maximum adult dose of 1 g) as oral suspension using dosing cups by trained CHWs. For children 1–11 months of age, dose will be determined by age.(8) For children 12– 59 months old, dose will be determined by height approximation as currently performed by Niger’s trachoma program.(9) Azithromycin will be prepared by Pfizer, Inc. (New York, NY, USA) and shipped directly to the study sites through the International Trachoma Initiative. Delayed or stopped treatment CSIs will receive usual care from the CHWs, which includes other community-based child health campaigns such as vitamin A supplementation, seasonal malaria chemoprevention, malnutrition screening, and immunization.

### Adverse Events

At each MDA, caregivers will be instructed to report adverse events to the CHWs if experienced within 14 days of treatment. The CHW will report adverse events to the CSI, which will then report to the study team. Non-serious adverse events will be reported to the study team in aggregate after each MDA, and serious adverse events (resulting in death, hospitalization, or those that are otherwise life-threatening) will be reported to the study team and medical monitors within 24 hours of receipt, and then to Pfizer, the Data and Safety Monitoring Committee (DSMC) and Institutional Review Boards according to the requirements of each. Children will receive care at the CSI or will be referred as needed.

### Outcome Assessments

*Mortality.* The primary mortality outcome will be defined as the under-5 mortality rate (U5MR, deaths per 1,000 births) as assessed by pregnancy history. A population-based census will be conducted in 50 randomly selected CSIs per region in Dosso, Tahoua, Maradi, and Zinder at baseline (25 per arm) and then approximately every 2 years.

Trained data collectors will visit each household in the study area to obtain verbal informed consent for participation. Household GPS coordinates and basic demographic data for the head of household and women of childbearing age (15-49 years old) will be collected. Full pregnancy histories will be conducted using procedures similar to those outlined in Demographic and Health Survey 8.(10, 11) Beginning with the first, each woman will be asked about each pregnancy she has experienced and pregnancy outcomes will be recorded for each. Current vital status, age, and sex will be recorded for all live births.

*Antimicrobial resistance.* AMR will be monitored in CSIs in the Maradi region that have not received prior azithromycin MDA for child survival. The CSIs will be randomly selected from among those selected for mortality monitoring in Maradi, with 20 randomly selected per arm. AMR will be monitored at baseline and then approximately every 2 years. At baseline and 2 years, this will include 20 CSIs from the treated and delayed arms. At 4 years, this will include 20 CSIs each from the continue, stop, and original delayed arms. Population-based and clinic-based monitoring will be conducted.

Population-based monitoring will occur in 3 randomly selected communities within each of the selected CSIs. Pregnancy history data will be used to identify eligible children 1-59 months of age, with 20 eligible children randomly selected for sample collections in each community. The study team will work with a local mobilizer to invite the selected group to a central location in the community for sample collection. Samples will be collected by trained personnel.

Nasopharyngeal swabs, rectal swabs, and dried blood spots will be collected from all selected children with verbal consent from their guardian using procedures detailed in prior studies.(12–14) CHWs will visit the study community to complete treatment for that round once sample collections have been completed.

Clinic-based monitoring will occur in all selected CSIs. Children 1-59 months of age presenting with symptoms of respiratory illness, diarrhea/dysentery, and for well-child visits will be considered for inclusion, with the goal of including 30 children in each group. The WHO Integrated Management of Childhood Illness flowchart will be used to identify children presenting with illness.(15) Nasopharyngeal swabs will be collected from children with respiratory illnesses. Rectal swabs will be collected from children with diarrhea symptoms. Both nasopharyngeal and rectal swabs will be collected from healthy children.

During collection, samples will be stored at ambient temperature or on ice at 4°C as required. At the end of each collection period, samples will be transported to a location near the community with facilities to store samples at −20°C or directly to the local study office each day. At the end of the collection period, all samples will be transferred to a central storage location in Niamey and stored at −20°C as required until processed in Niger or shipped to UCSF for processing.

*Implementation outcomes.* The RE-AIM framework was used to define implementation outcomes for evaluation (Table 2).(16, 17) The following additional data collection activities will take place to measure these outcomes:

- **Coverage and Coverage Equity.** Treatment coverage will be assessed in several ways. CHWs will record key information about each MDA on paper data collection forms, including the the number of treated children, adverse events experienced, and the number of days required for the distribution. Administrative coverage will be estimated using current estimates of the under-5 population from CSIs as the denominator and the number of children treated in each MDA as the numerator according to the CHW data collection form and will be used for official coverage reporting for all CSIs. Census coverage will be calculated using estimated eligible population denominators available from the pregnancy histories conducted in the monitoring CSIS. Administrative and census coverage will be compared to determine the validity of the administrative coverage estimates.

**Table 2.**
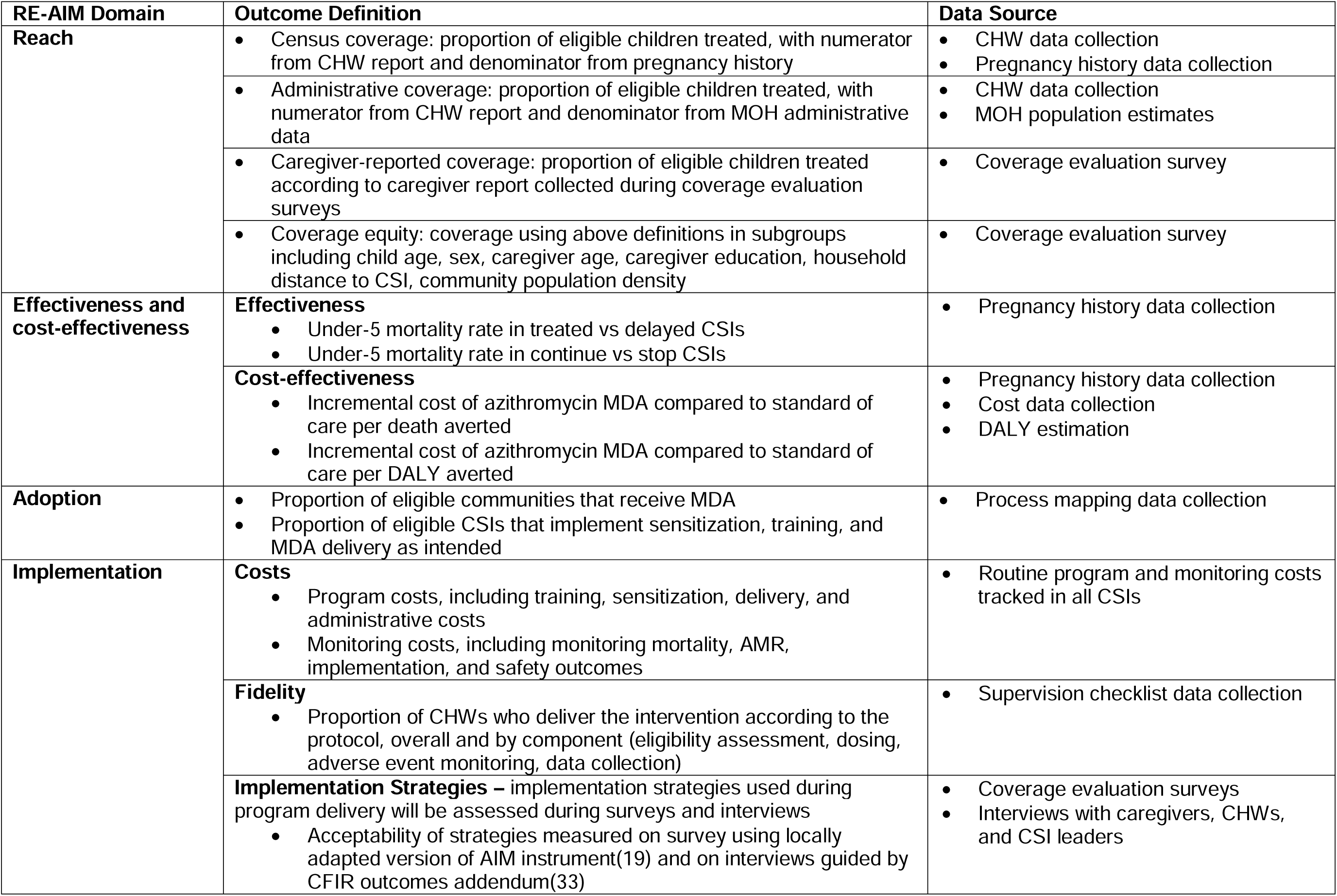
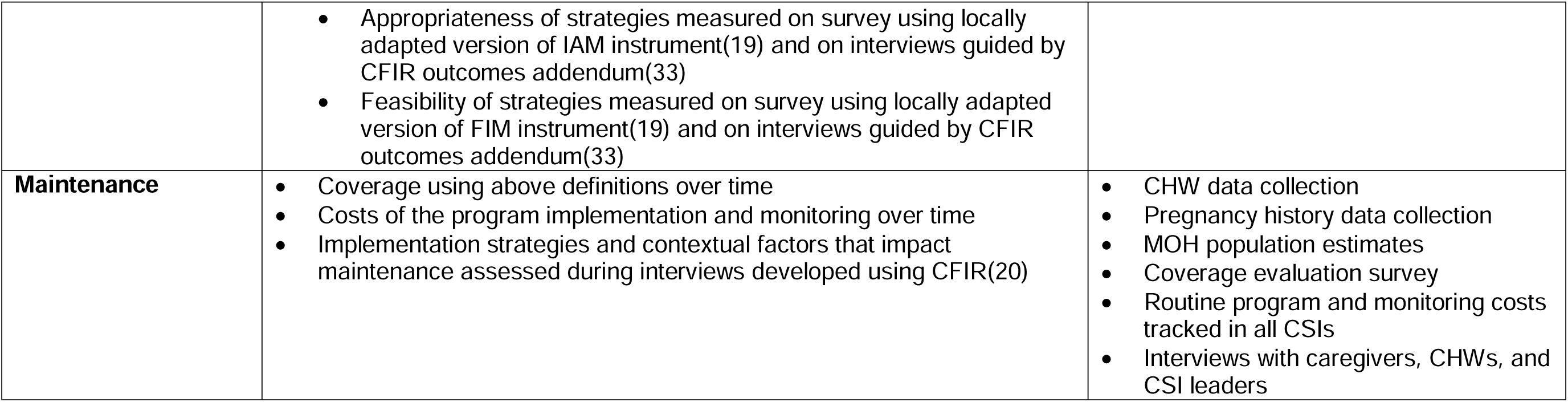
RE-AIM framework applied to AVENIR II outcomes.

| **RE-AIM Domain** | **Outcome Definition** | **Data Source** |
| --- | --- | --- |
| **Reach** | Census coverage: proportion of eligible children treated, with numerator from CHW report and denominator from pregnancy history | CHW data collection<br>Pregnancy history data collection |
|  | Administrative coverage: proportion of eligible children treated, with numerator from CHW report and denominator from MOH administrative data | CHW data collection<br>MOH population estimates |
|  | Caregiver-reported coverage: proportion of eligible children treated according to caregiver report collected during coverage evaluation surveys | Coverage evaluation survey |
|  | Coverage equity: coverage using above definitions in subgroups including child age, sex, caregiver age, caregiver education, household distance to CSI, community population density | Coverage evaluation survey |
| **Effectiveness and cost-effectiveness** | **Effectiveness**<br>Under-5 mortality rate in treated vs delayed CSIs<br>Under-5 mortality rate in continue vs stop CSIs | Pregnancy history data collection |
|  | **Cost-effectiveness**<br>Incremental cost of azithromycin MDA compared to standard of care per death averted<br>Incremental cost of azithromycin MDA compared to standard of care per DALY averted | Pregnancy history data collection<br>Cost data collection<br>DALY estimation |
| **Adoption** | Proportion of eligible communities that receive MDA<br>Proportion of eligible CSIs that implement sensitization, training, and MDA delivery as intended | Process mapping data collection |
| **Implementation** | **Costs**<br>Program costs, including training, sensitization, delivery, and administrative costs<br>Monitoring costs, including monitoring mortality, AMR, implementation, and safety outcomes | Routine program and monitoring costs tracked in all CSIs |
|  | **Fidelity**<br>Proportion of CHWs who deliver the intervention according to the protocol, overall and by component (eligibility assessment, dosing, adverse event monitoring, data collection) | Supervision checklist data collection |
|  | **Implementation Strategies** – implementation strategies used during program delivery will be assessed during surveys and interviews<br>Acceptability of strategies measured on survey using locally adapted version of AIM instrument(19) and on interviews guided by CFIR outcomes addendum(33) | Coverage evaluation surveys<br>Interviews with caregivers, CHWs, and CSI leaders |
|  | • Appropriateness of strategies measured on survey using locally adapted version of IAM instrument(19) and on interviews guided by CFIR outcomes addendum(33)<br>• Feasibility of strategies measured on survey using locally adapted version of FIM instrument(19) and on interviews guided by CFIR outcomes addendum(33) |  |
| **Maintenance** | • Coverage using above definitions over time<br>• Costs of the program implementation and monitoring over time<br>• Implementation strategies and contextual factors that impact maintenance assessed during interviews developed using CFIR(20) | • CHW data collection<br>• Pregnancy history data collection<br>• MOH population estimates<br>• Coverage evaluation survey<br>• Routine program and monitoring costs tracked in all CSIs<br>• Interviews with caregivers, CHWs, and CSI leaders |

Coverage evaluation surveys will be conducted in a randomly selected subset of CSIs to validate coverage estimates using caregiver self-report, to identify subgroups with low coverage to evaluate coverage equity, to understand reasons for low coverage, and to compare outcomes of different implementation strategies. Survey instruments will be developed based on standardized questionnaires used for neglected tropical disease program monitoring according to WHO guidance,(18) and implementation strategies will be assessed using versions of the Acceptability of Intervention Measure, Intervention Appropriateness Measure, and Feasibility of Intervention Measures that have been modified to ensure cultural appropriateness.(19) Trained data collectors fluent in local languages will visit every household in selected CSIs and conduct the survey among caregivers of children eligible for the program after obtaining verbal informed consent. Surveys will take place within 1 week of the most recent MDA.

In-depth interviews will be conducted after 2 and 4 years in a random sample of caregivers stratified by participation in the program as well as a random sample of CHWs and CSI leaders to identify barriers and facilitators to participation and implementation along with implementation strategies to improve uptake. Interview guides will be developed using the Consolidated Framework for Implementation Research (CFIR).(20) Trained local interviewers fluent in relevant languages will conduct semi-structured interviews in private locations after obtaining verbal informed consent.

Interview recordings and notes will be used for rapid qualitative analysis to immediately summarize each interview according to pre-defined domains, which will then be distilled into a matrix organized by domain and interviewee for analysis.(21) The first 5 interviews will be summarized by the group of interviewers as part of training to achieve consensus on the approach.

- **Process Mapping.** Process mapping will be used to identify the flow of inputs required to achieve high MDA coverage along with information on which aspects of the intervention are adaptable and which are key determinants of high coverage. Following methods used in other MDA settings,(22) we will use routine workflow tracking in the same CSIs selected for coverage evaluation surveys to record data on key activities during each round of MDA, including drug supply chain, training, sensitization, and the MDA itself. After 2 years, we will use coincidence analysis to identify any necessary and/or sufficient combinations of intervention activities that resulted in high MDA coverage.(23)
- **Costs and Cost-effectiveness.** Routine expenses will be tracked in all CSIs to monitor overall program costs, costs of program components (training, sensitization, delivery, administrative), costs per district, and costs per dose delivered. Incremental cost-effectiveness of the intervention compared to standard of care will be estimated using program costs, with effectiveness outcomes including measured mortality and estimated DALYs to calculate cost per death averted and cost per DALY averted.
- **Fidelity.** We will monitor CHW implementation using two methods: local CSI leaders will supervise CHWs in nearby communities according to their standard practice, and the study team will monitor fidelity to the protocol using standardized checklists in the same random sample selected for coverage evaluation surveys. These checks ensure the proper execution of program procedures, including eligibility assessment, dosing, adverse event monitoring, and data collection.

*Morbidity.* Clinic visits will be monitored through passive surveillance at all CSIs. CSIs routinely record details of all clinic visits in log books which are aggregated into counts of specific diagnoses and treatments by CSI and entered into Niger’s DHIS2 database. All infectious-cause visits and cause-specific visits for diagnoses of respiratory infection, diarrhea, and malaria will be extracted among children under 5. Records will be regularly reviewed for eligible entries and data including child age, community of residence, date of visit, diagnosis at visit, and treatment given will be extracted for all CSIs. Diagnoses of respiratory infection, diarrheal disease, and malaria at the CSIs are determined using the Integrated Management of Childhood Illness flowchart.(15)

### Data Collection, Management, and Monitoring

All study personnel who assess outcomes will attend two-day training sessions prior to conducting study procedures. Training will include the operation of the mobile device where relevant, data collection through the mobile application or on paper, and the study protocol and procedures. Where possible, data will be collected electronically using a custom-designed mobile census application (CommCare, Dimagi) on mobile devices and uploaded regularly to a secure, cloud-based server. Where possible, data will be uploaded in real-time using the cellular network, or weekly or monthly at a central site with strong Wi-Fi or cellular connection. Mobile devices will be encrypted and accessed via unique usernames and passwords. Mobile devices will be stored in locked offices with electricity for charging. Data on intervention administration will be recorded by community health workers on paper forms and later entered electronically.

Validation will be used in the development of the data collection forms to reduce inconsistencies and invalid entries. Data management teams in Niger and at UCSF will monitor data collection in real-time daily. Progress and quality control reports will be prepared and distributed to investigators weekly. Study participants will be assigned unique identification numbers that do not contain identifying information, which will be used to link data across databases and will be the only identifier shared in de-identified datasets.

### Study Oversight

*Niger MOH committee.* Prior to the start of study activities, an MOH-based committee was established to approve study design and protocols and oversee implementation of all activities. The committee includes MOH members with expertise in pediatrics, antimicrobial resistance, and community health interventions. Annual meetings will be held to review progress reports and make decisions about study continuation, recommendations from the DSMC, adding/dropping arms, and program implementation.

*Institutional Review Boards.* The Comité National Éthique pour la Recherche en Santé in Niger (14/2024/CNERS) and University of California San Francisco Institutional Review Board (23–39839) have given ethical approval for the trial and will review the trial annually.

*DSMC.* A DSMC was empaneled before the study began to provide independent oversight of data quality and participant safety. The DSMC includes independent experts with collective expertise in bioethics, biostatistics, antimicrobial resistance, and pediatric infectious disease. The DSMC will meet at least once per year to review study progress and adverse events, with ad hoc meetings convened as necessary. The DSMC will recommend modifications to the protocol as necessary.

In addition to the groups mentioned above, study investigators from UCSF and in Niger will monitor study conduct regularly through data monitoring as described previously in addition to regular site visits to ensure adherence to the study protocol. Two Medical Monitors with expertise in pediatrics and infectious disease will review the protocol before the trial begins and will review serious adverse events during the trial. The UCSF and Niger PIs will determine eligibility for authorship based on contributions to each manuscript and the related activities.

### Sample size and power

For the overall program, we estimate that approximately 3 million children will be eligible to receive the intervention in approximately 1300 CSIs in 7 regions in Niger. We currently plan to monitor the primary mortality and AMR outcomes in 200 CSIs, evenly stratified across the 4 regions starting the program (Dosso, Tahoua, Maradi, Zinder). Calculations are presented for the first trial comparing treated to delayed arms and are assumed to apply to the second trial comparing continue to stop treatment arms as well.

*Under 5 Mortality Rate (U5MR).* We estimate that monitoring a total sample of 200 CSIs, evenly split between MDA and delayed, would have 80% power to detect a reduction in the U5MR of 6.6 per 1000 live births. At 90% power, the reduction in U5MR is approximately 7.7 per 1000 live births. These estimates are based on a sample size equation for a t-test of cluster-level means. We assumed an alpha of 0.05 and a standard deviation of 16.6 in the CSI-level U5MR, which was estimated from the MORDOR Niger trial 5-year birth history substudy.(24)

*AMR.* We plan to monitor 20 randomly selected CSIs per arm and 3 randomly selected communities per CSI, giving a total of 60 communities per arm. Within each community, 15 children will be randomly selected from the prior census to receive sample collections.

- <u>Nasopharyngeal samples</u>. A total of 120 communities (60 per arm) will provide approximately 80% power to detect a 7% absolute increase in the prevalence of macrolide-resistant pneumococcus isolates different between arms. This assumes a 10% baseline prevalence of macrolide resistance, a community-level ICC of 0.045 estimated from MORDOR I 24-month data, and an alpha of 0.05.(12)
- <u>Rectal samples:</u> A total of 120 communities (60 per arm) will allow 80% power to detect a 1.4-fold increase in community mean normalized read number of genetic determinants of resistance to macrolides in rectal samples. This assumes an estimated ICC of 0.02 from the MORDOR I 24-month data, a standard deviation of 3.1 (on the logarithmic scale), and an alpha of 0.05.(12)

*Implementation Outcomes.* Sample size calculations were conducted for the coverage evaluation surveys. The surveys will be conducted in a randomly selected subset of 3 CSIs per region among the CSIs after each MDA. Within each CSI, 7 communities will be randomly selected to participate in the survey. We estimate that including approximately 2,100 households per region will allow us to estimate caregiver-reported coverage of 70% ± 5% at the region level, assuming an average of 100 households per community-based on AVENIR I, a design effect of 4 based on guidance from the WHO, an alpha of 0.05, and non-response of 10%.(3, 18)

### Statistical considerations

Statistical methods are described in detail for the primary mortality and AMR outcomes which will involve intention to treat analyses.

*Primary outcome definitions.* The primary mortality outcomes are defined as:

- Under-5 mortality rate (U5MR, deaths per 1,000 live births) assessed by pregnancy history at 2 years from the first treatment distribution, comparing the intervention and delayed arms
- Under-5 mortality rate (U5MR, deaths per 1,000 live births) assessed by pregnancy history at 4 years, comparing the continue and stop arms

We will calculate the U5MR at the CSI level as the cumulative probability of mortality by age 60 months using a synthetic cohort approach from pregnancy history records.(25)

The primary AMR outcomes are defined as:

- Prevalence of macrolide-resistant pneumococcus from nasopharyngeal swabs in children 1–59 months old after 2 years of distributions, comparing the intervention and delayed arms
- Prevalence of macrolide-resistant pneumococcus from nasopharyngeal swabs in children 1–59 months old after 4 years of distributions, comparing the continue and stop arms
- Load of genetic determinants of resistance to macrolides from rectal swabs in children 1–59 months old after 2 years of distributions, comparing the intervention and delayed arms
- Load of genetic determinants of resistance to macrolides from rectal swabs in children 1–59 months old after 4 years of distributions, comparing the continue and stop arms

*Analysis.* The primary analysis for U5MR will be to compare the proportion of children who die before age 60 months between the two arms. The prior likelihood for mortality will be estimated using cluster-level data from the AVENIR I trial, using methods similar to the likelihood estimation.(3) We will model CSI-level U5MR using a generalized linear regression model with terms for treatment arm and baseline U5MR. CSIs should be of similar size, but if we find there are large differences in size then we will consider weighting the regression by the number of measurements in each CSI. We will estimate the likelihood of the data from the fitted regression model. Note that the MOH committee and DSMC will review and approve the prior used past the second year of the trial, with options including incorporation of new results outside of AVENIR I and II, as well as down-weighting the prior from AVENIR I over time.

To estimate the posterior, beginning with the maximum likelihood estimate of all parameters obtained by the generalized linear model, we will choose particular values of the treatment effect β. For each such value, we maximize the likelihood with respect to the other parameters, yielding the profile likelihood function with respect to the single parameter β. We will multiply the profile likelihood by the prior to obtain the posterior distribution, normalized to ensure the posterior likelihood sums to 1. We will derive Bayesian credible intervals from the posterior derived in this way. Specifically, we will report the 95% central coverage interval and the estimated median. We will also estimate the probability of the null hypothesis under the posterior likelihood, and its complement (the probability MDA reduces the U5MR). Subgroup analyses for U5MR include region, child age, child sex, and community distance to the CSI. Note that for analyses of the second trial comparing the continue vs stop arms, the first 6 months will be excluded from analyses to allow for a washout period to account for lingering effects from prior treatment.

The Bayesian credible intervals of the U5MR will also be used to compare the measured mortality rate against the Sustainable Development Goal (SDG) targets (a secondary outcome). We will estimate the probability that the U5MR is at or below the SDG target using the area of the posterior distribution that falls at or below the SDG target.

The primary analysis for resistance outcomes from community-based collection of nasopharyngeal (NP) swabs will be to compare the proportion of macrolide-resistant pneumococcal isolates between the two arms. If the laboratory is not able to grow pneumococcal isolates in at least 30% of the samples we will use DNA testing instead.

The primary analysis for the community-based collection of rectal swabs will compare the community-level load of macrolide-resistant genetic determinants between the two arms. For both sets of samples, secondary analyses examining resistance in other classes of antibiotics, other age groups, and among individual antibiotic resistance genes will be conducted similarly.

*Missing data.* Monitoring communities that are not measured at year 2 or year 4 will be reported, but their measurements in that phase will not contribute to the analysis. If the number of communities lost exceeds 15% (30 of 200) we will conduct a sensitivity analysis that uses inverse probability of censoring weights to correct for potential bias due to potentially informative censoring (assuming outcomes are missing at random, conditional on covariates). Inverse probability weights will be estimated using logistic regression and all community-level characteristics measured at baseline will be considered in the model.

*Interim analysis and stopping guidelines.* There are no planned interim analyses or guidelines for early stopping at this time. Interventions may be stopped or changed at the discretion of the Niger MOH committee in discussion with the study team and the DSMC.

## DISCUSSION

Weighing the mortality benefit against the AMR risk has proven a consistent challenge in discussions of programmatic implementation of azithromycin MDA. The WHO decided that the more proximal child survival benefits outweighed the uncertain distal AMR risks,(4) but others have expressed alarm at the concept of the intervention in this era of antimicrobial stewardship and believe it should not be delivered at all.(26, 27) Some groups have concerns over “ethical double standards,” where azithromycin MDA is criticized by those in high-income settings that do not face a high mortality burden.(28, 29) When compared to other widely accepted uses of antimicrobials, azithromycin MDA has been shown to fall well within a justifiable range for metrics like number needed to treat to save one life and risk-benefit trade-off.(30) Regardless, nearly all agree that any implementation of azithromycin MDA should be accompanied by close monitoring of impact on AMR, and that the countries and communities considering implementation of the program should lead any decision-making. Indeed, the Niger MOH determined that concerns over AMR warranted a program rollout that included a randomized design to allow for causal inference about the effectiveness of azithromycin MDA on mortality and the impact on resistance.

Here we present a flexible design that allows for the evaluation not only of the azithromycin MDA intervention but others as well. As the epidemiologic landscape shifts over time, the effectiveness of child survival programs and priorities for community health may change. Azithromycin MDA for child survival may no longer be warranted as mortality declines or if the intervention contributes to detrimental increases in AMR. However, attributing effects to the program is challenging without a rigorous monitoring design. For example, longitudinal monitoring of AMR could show consistent increases over time due to antibiotic use outside of the program, even if azithromycin MDA remains effective in reducing mortality with only small and transient impacts on resistance. Thus, a monitoring design that allows for causal attribution is warranted. Moreover, despite the increasingly popularity of and recent methodological advancements in adaptive and platform trials, use of such methods in global health settings remains rare.(31) Among other benefits, adaptive designs can allow for more ethical randomization through methods like response adaptive randomization, enabling preferential treatment with more effective interventions over time.(32) Platform trials leverage the upfront investments made in trial infrastructure by examining multiple arms, dropping ineffective arms, and adding arms to adapt to new data and shifting priorities.(6) The use of an adaptive platform design allows us to monitor the impact of azithromycin MDA on mortality and AMR over time, enabling both the ability to determine whether to stop or continue the program as well as to evaluate other interventions as priorities shift.

Limitations of this design include the lack of a placebo control, which increases the potential for bias. However, as multiple prior placebo-controlled trials have demonstrated the efficacy of this intervention and eligible countries are ready to move to large scale program implementation, placebo is no longer warranted. In addition, eligible communities in Niger receive multiple community-based health campaigns from the same CHWs each year, limiting concerns of a Hawthorne effect. The design does allow for masking of all outcome assessors, reducing the risk of bias. Another potential limitation involves statistical power. As mortality is declining over time outside of this intervention, and heterogeneity in outcomes may be greater compared to prior studies given the large scale of the implementation, the trial could have less power to detect effects than anticipated based on current assumptions. Power calculations for this trial used data from the AVENIR I trial, which includes the most recent population-based mortality data available in Niger from multiple regions to address this concern. Finally, the shift from a controlled trial setting to a real-world program will involve changes in the implementation of the program itself which could impact the ability to detect effects, as there will likely be more variation in program delivery across settings and the program may see lower and more heterogenous MDA coverage than the prior trials. Close monitoring of coverage and factors contributing to coverage heterogeneity were included in the trial design to allow for such evaluation.

AVENIR II will provide evidence on mortality, AMR, implementation, and safety outcomes after large-scale programmatic delivery of azithromycin MDA for child survival in Niger. Results from this trial will support program-related decision-making related in Niger and elsewhere. The adaptive platform design provides a flexible approach to allow for this monitoring and to evaluate other community interventions over time.

## TRIAL STATUS

This manuscript describing the AVENIR II protocol was developed based on the official study protocol version 12 which was last revised on 26 September 2024. Recruitment began in June 2024 and is expected to be completed May 2028.

## LIST OF ABBREVIATIONS

AMR: Antimicrobial Resistance
CDC: Center for Disease Control
CSI: Centre de Santé Integré
CFIR: Consolidated Framework for Implementation Research
IRB: Institutional Review Board
MDA: Mass Drug Administration
MOH: Ministry of Health
RE-AIM: Reach, Effectiveness, Adoption, Implementation, Maintenance
SAP: Statistical Analysis Plan
SDG: Sustainable Development Goal
UCSF: University of California San Francisco
U5MR: Under 5 Mortality Rate
WHO: World Health Organization

## DECLARATIONS

### Ethics approval and consent to participate

The University of California San Francisco Institutional Review Board has given ethical approval for the trial and will review the trial annually (23–39839). The Comité National Éthique pour la Recherche en Santé in Niger granted ethical approval before study activities began (DELIBERATION No. 14/2024/CNERS). Verbal informed consent will be obtained from the head of household for participation in the census and from caregiver/guardians for treatment administration and outcome data collection. Verbal consent will be obtained from regional, district, and community leaders before recruiting any participants into the study. There is no financial cost to the participant and there is no reimbursement for overall participation in this study.

### Consent for publication

No data have been included in this study protocol. No details, images, or videos relating to individuals have been included. The authors will provide the verbal consent scripts used in this study upon request.

### Availability of data and materials

Trial results will be presented at local, national and international meetings and submitted to peer-reviewed journals for publication. De-identified trial data will be made publicly available after publication of primary outcomes.

## Authors’ contributions

*All authors contributed to the design of the trial. KSO, EL, BFA, and TML prepared the initial draft of the manuscript, and all authors provided critical review and revisions of the manuscript. All authors read and approved the final manuscript*.

## Data Availability

De-identified trial data will be made publicly available at OSF after publication of primary outcomes.

## Acknowledgements

The authors would like to acknowledge the AVENIR II study group and partners for their many contributions to this work: ***Centre de recherche et interventions en santé publique, Niamey, Niger*** (Moustapha Mamane Abarchi, Fati Adamou, Nana Hadiza Bakalmale Adamou, Ahmed M. Arzika, Ismael Mamane Bello, Annatou Boubacar, Mariama Boubacar, Ousseini Soumana Boubacar, Abdouramane Gado Djibo, Abass Harouna Dodo, Abdell Nasser Harouna Dodo, Mahamadoul-Nasser Mamane Galo, Nana Fatchima Mamane Gallo, Maman Laminou Maliki Haroun, Ahmed Harouna, Lalla Hamet Hassan, Karamatoulaye Hamadou Ide, Alio Karamba, Abdoul Lahi Ibrahim Mahamane, Ramatou Maliki, Barhamou Oumarou Nomao, Farissatou Oumarou, Zabayrou Chaibou Oumarou, Mahamadou Djamilou Sani Ousmane, Nana Mardiyatou Ibrahim Salifou, Aichatou Bawa Salissou, Mariama Tiemogo Souley, Almoustapha Hamma Souna) ; ***Centre de recherche médical et sanitaire, Niamey, Niger*** (Inoussa Dodo Abdoulaye, Zainabou Abdoulkarim Benoit, Mahamadou Dobzanga, Sani Haladou, Maman Koraou Halimatou, IIdrissa Hamadou, Sani Ousmane, Sabo Haoua Seini, Amadou Moussa Soussou, Zara Tahirou, Nana Bassira Tidjani) ; ***Centre de santé communautaire, Niamey, Niger*** *(Amadou Zakari)* ; ***Direction des Statistiques, Niamey, Niger*** (Aida Mounkala, Salamatou Maiga, Mahamadou Yahaya) ; ***Centre de santé de la mère et de l’enfant, Dosso, Niger*** (Amina Seyfoulaye) ; ***International Trachoma Initiative*** (David Addiss, Genevieve Emidy, PJ Hooper, Carla Johnson, Kristin Renneker, Moumine Yaro) ; ***Pfizer, New York, NY, USA*** (Todd D. Hatajik, Julie Jenson, Chiao-Chin Lin); ***Programme national de santé oculaire, Niamey, Niger*** (Amza Abdou, Zakari Adamou, Hassane Amadou, Abdrahamane Bachabi,Hamidou Amadou Bagna, Yacouba Baraze, Namewa Boubacar, Hassane Khady Diop, Almou Ibrahim, Baido Nassirou, Laouali Lamine ) ; ***University of California, San Francisco, San Francisco, CA, USA*** (Benjamin Arnold, Carolyn Brandt, Cindi Chen, Angela Cheng, Thuy Doan, Jeremy D Keenan, Elodie Lebas, Thomas M Lietman, Zijun Liu, Kieran S O’Brien, Catherine E Oldenburg, Brittany Peterson, Andrea Picariello, Travis C Porco, Kevin Ouimette, Lina Zhong, Zhaoxia Zhou); ***University of Maryland School of Medicine, Baltimore, USA*** (Meagan Fitzpatrick, Karen Kotloff)

In addition, the authors would like to thank the members of the Data and Safety Monitoring Committee (*Emory University, Task Force for Global Health –* David Addiss (chair); *Niger Ministry of Health* – Mourtala Assao; *St. George’s University of London* – Julia Bielicki, *Retired from CDC* – Allen Hightower, *Children’s Hospital of Colorado* – Brian Jackson, *Hopital National de Niamey, Niger* – Moumouni Kamaye, *Emory University –* Jim Lavery, *University of Melbourne* – Fiona Russell, *Hopital Amirou* Boubacar Diallo*, Niamey, Niger* – Abdourahamane Yacouba) and all members of the Niger MOH Committee.

